# Has the age distribution of hospitalized Covid-19 patients changed in Brazil?

**DOI:** 10.1101/2021.03.30.21254650

**Authors:** Carla Lourenço Tavares de Andrade, Sheyla Maria Lemos Lima, Monica Martins, Claudia Cristina de Aguiar Pereira, Margareth Crisóstomo Portela

**Author notes:** Corresponding author: Margareth Crisóstomo Portela.

## Abstract

The aim of this study was to compare the age profile of hospitalized Covid-19 patients during the first year of the pandemic, as well as hospital mortality and use of ICUs, by age group, in large geographic regions of Brazil. We used data from the Influenza Epidemiological Surveillance Information System for patients who presented the first symptoms of the disease between the epidemiological weeks 8 of 2020 and 7 of 2021, which were divided into three periods.

779,257 records of patients hospitalized by Covid-19 were obtained. Of this total, 720,363 (92.4%) referred to discharged hospitalizations, considered in the analysis of ICU use and death. Among 244,611 hospitalizations (34.0%) with indication for use of ICU, 190,833 allowed the calculation of the time in ICU. There was variation in the age profile of hospitalized patients between the three periods, but there was no evidence in favor of the hypothesis of an increase, in the last period, in the participation of adults between 18 and 50 years old in hospitalizations by Covid-19. A differentiated increase in the mortality of young adults in the North suggests the possibility of greater severity of the P1 variant in this population. The results also show that the participation of young adults in hospitalizations and hospital deaths was never negligible and is related to hospital mortality rates close to or above 10%.

The Covid-19 “youthening” phenomenon in Brazil is based on the country’s own sociodemographic and economic characteristics and may have been strengthened by the increasing circulation of viral variants. It is important to continue monitoring its progression and effects.

## Introduction

Brazil ended the first year of the Covid-19 pandemic in a scenario of increasing number of cases, with almost simultaneous growth in the rates of incidence and mortality due to the disease and high levels of occupancy of intensive care beds (ICU) in the 26 states and the Federal District, often greater than 90%^1^. During the first year, regions of the country were most affected at different times, with maintenance of the daily moving average at more than a thousand deaths between June and August 20202. There was, then, a drop in the average of deaths until the beginning of November, but, in January 2021, the moving average again exceeded one thousand deaths, reaching, in March, more than two thousand daily deaths, a figure never seen before^2^. Currently, Brazil is the second country with the highest absolute number of cases (11,998,233) and deaths (294,042)^3^, and accounts for more than 1/4 of the registered deaths globally, with only 2.7% of the world population^4^.

After a year of pandemic, the most critical moment of the health and care crisis is experienced^1,5^. The low adherence of the population to the pandemic control and prevention measures, aggravated during the last months of 2020, and the appearance of new variants of the SARS-CoV-2 virus, which are more transmissible than the original version^6^, are plausible explanations for the high growth of cases and deaths.

In this context, there is frequent conjecture about a possible change in the age profile of patients hospitalized by Covid-19 in the country, with younger people presenting more severe conditions of the disease, higher mortality and more prolonged use of ICU beds^7^. The aim of this study was to compare, during the first year of the pandemic, the age profile of patients hospitalized by Covid-19, as well as hospital mortality and use of ICUs, by age group, in large geographic regions of Brazil.

## Methods

This study used data from the Influenza Epidemiological Surveillance Information System (SIVEP-Gripe) from DATASUS, Ministry of Health, which is public and unrestricted. The system is nationwide and includes compulsory notifications about severe acute respiratory syndrome, including demographic and clinical variables, related to the use of outpatient and hospital services and the outcome of the case. Hospital data include the use and dates of entry and exit from the ICU. The data were accessed on the website https://opendatasus.saude.gov.br on March 17, 2021. Of the records from 2020 and 2021, those with a final classification of ‘SRAG by COVID-19’ that involved hospitalization were considered.

For each region of the country, the age profiles of hospitalized patients were compared, and in each age group, the occurrence of hospital mortality and ICU use, as well as the time (days) in the ICU, considering three periods during the first year of the pandemic. Cases were included for those with first symptoms occurring between the eighth epidemiological week of 2020 and the seventh epidemiological week of 2021. The first period included 18 epidemiological weeks, between 16 February and 20 June 2020; the second included 18 epidemiological weeks, between 21 June and 24 October 2020; and the third included 17 epidemiological weeks, between October 25, 2020 and February 20, 2021. The time frame until the seventh epidemiological week of 2021 allowed us to examine the first year of the pandemic, mitigating the problem of dealing with the underreporting of cases in other available weeks and excessive loss of unfinished records.

For comparisons of the age profiles of hospitalized patients, between periods, all records were counted in the calculation of absolute and relative frequencies of hospitalizations, using the chi-square test. The comparisons of ICU use and the occurrence of death by age group considered only the records with discharge information. For comparing the ICU length of stay, the closed records that informed the date of admission and discharge from ICU. In the absence of the date of exit and occurrence of death, it was assumed that the date of exit was the date of death. In all three cases, the Kruskal-Wallis non-parametric test was applied.

## Results

779,257 records of patients hospitalized by Covid-19 were obtained. Of this total, 720,363 (92.4%) had data (type and / or date) of the patient’s evolution or had been closed, making up the set of observations used in the analysis of ICU use and death. Among these, 244,611 (34.0%) contained an indication of ICU use, and 190,833 had data that allowed the calculation of ICU length of stay.

Table 1 shows that there was variation in the age profile of hospitalized patients between the three periods considered in the regions of the country, however it does not provide evidence in favor of the hypothesis of an increase, in the last period, of hospitalizations of people between 18 and 50 years, compared to previous periods of the pandemic. In the North and Northeast regions, the proportion of individuals aged between 18 and 50 years old, in the last period, approached those observed in the first period, when they were also substantially affected by the pandemic. In the Southeast and South, there was a consistent reduction in these age groups over the periods. In the Central West region, which was mostly affected only after the second period, the participation of adults between 18 and 50 years old in hospitalizations was higher in the first period, decreasing in the second and remaining relatively close to the second in the third.

**Table 1.** **Comparison of the distribution of Covid-19 hospitalizations (N = 779,257) by age groups among three periods of the pandemic’s first year in the regions of Brazil**

| Region/ Age<br>group (years) | Epidemiologic weeks | | | | | | $\chi^2$<br>(p) |
| --- | --- | --- | --- | --- | --- | --- | --- |
|  | 08/2020 – 25/2020 |  | 26/2020 – 43/2020 |  | 44/2020 – 07/2021 |  |  |
|  | (16 Feb 2020 – 20 Jun 2020) |  | (21 Jun 2020 – 24 Oct 2020) |  | (25 Oct 2020 – 20 Feb 2021) |  |  |
|  | N | % | N | % | N | % |  |
| North |  |  |  |  |  |  | <0.0001 |
| 0-5 | 351 | 1.5 | 793 | 4.8 | 744 | 3.0 |  |
| 6-17 | 341 | 1.4 | 499 | 3.1 | 275 | 1.1 |  |
| 18-30 | 1374 | 5.8 | 1310 | 8.0 | 1245 | 5.0 |  |
| 31-40 | 2646 | 11.2 | 1765 | 10.8 | 2749 | 11.1 |  |
| 41-50 | 3476 | 14.8 | 2178 | 13.3 | 3735 | 15.0 |  |
| 51-60 | 3990 | 17.0 | 2614 | 16.0 | 4491 | 18.1 |  |
| 61-70 | 4751 | 20.2 | 2968 | 18.1 | 5262 | 21.2 |  |
| 71-80 | 3990 | 17.0 | 2594 | 15.8 | 3916 | 15.8 |  |
| >80 | 2622 | 11.1 | 1662 | 10.1 | 2416 | 9.7 |  |
| Total | 23541 | 100.0 | 16383 | 100.0 | 24833 | 100.0 |  |
| Northeast |  |  |  |  |  |  | <0.0001 |
| 0-5 | 804 | 1.4 | 980 | 2.3 | 782 | 1.9 |  |
| 6-17 | 530 | 0.9 | 713 | 1.7 | 378 | 0.9 |  |
| 18-30 | 2806 | 4.9 | 1981 | 4.6 | 1683 | 4.0 |  |
| 31-40 | 5539 | 9.7 | 3659 | 8.6 | 3993 | 9.6 |  |
| 41-50 | 7527 | 13.2 | 4887 | 11.4 | 5434 | 13.1 |  |
| 51-60 | 9744 | 17.1 | 6711 | 15.7 | 7342 | 17.6 |  |
| 61-70 | 11238 | 19.8 | 8206 | 19.2 | 8468 | 20.3 |  |
| 71-80 | 10646 | 18.7 | 8437 | 19.8 | 7609 | 18.3 |  |
| >80 | 8049 | 14.2 | 7128 | 16.7 | 5935 | 14.3 |  |
| Total | 56883 | 100.0 | 42702 | 100.0 | 41624 | 100.0 |  |
| Southeast |  |  |  |  |  |  | <0.0001 |
| 0-5 | 948 | 0.9 | 1099 | 0.9 | 1185 | 0.8 |  |
| 6-17 | 573 | 0.5 | 881 | 0.7 | 653 | 0.4 |  |
| 18-30 | 4541 | 4.1 | 4648 | 3.7 | 4611 | 3.1 |  |
| 31-40 | 11962 | 10.8 | 11294 | 9.0 | 12509 | 8.5 |  |
| 41-50 | 17410 | 15.8 | 16986 | 13.5 | 19636 | 13.3 |  |
| 51-60 | 21627 | 19.6 | 23316 | 18.5 | 27908 | 18.9 |  |
| 61-70 | 22341 | 20.2 | 27419 | 21.8 | 33711 | 22.8 |  |
| 71-80 | 17400 | 15.8 | 22707 | 18.0 | 27429 | 18.5 |  |
| >80 | 13559 | 12.3 | 17551 | 13.9 | 20326 | 13.7 |  |
| Total | 110361 | 100.0 | 125901 | 100.0 | 147968 | 100.0 |  |
| South |  |  |  |  |  |  | <0.0001 |
| 0-5 | 64 | 0.7 | 301 | 0.7 | 317 | 0.5 |  |
| 6-17 | 41 | 0.4 | 232 | 0.5 | 225 | 0.3 |  |
| 18-30 | 570 | 5.8 | 1725 | 4.0 | 2160 | 3.3 |  |
| 31-40 | 1166 | 11.8 | 4083 | 9.4 | 5678 | 8.7 |  |
| 41-50 | 1722 | 17.5 | 6127 | 14.0 | 9075 | 13.8 |  |
| 51-60 | 2048 | 20.8 | 8648 | 19.8 | 12952 | 19.8 |  |
| 61-70 | 1937 | 19.7 | 9740 | 22.3 | 15279 | 23.3 |  |
| 71-80 | 1408 | 14.3 | 7528 | 17.2 | 12096 | 18.5 |  |
| >80 | 886 | 9.0 | 5290 | 12.1 | 7719 | 11.8 |  |
| Total | 9842 | 100.0 | 43674 | 100.0 | 65501 | 100.0 |  |
| Central West |  |  |  |  |  |  | <0.0001 |
| 0-5 | 65 | 0.7 | 368 | 1.0 | 146 | 0.6 |  |
| 6-17 | 49 | 0.5 | 232 | 0.6 | 119 | 0.5 |  |
| 18-30 | 506 | 5.5 | 1562 | 4.2 | 1032 | 4.3 |  |
| 31-40 | 1272 | 13.9 | 3762 | 10.2 | 2528 | 10.6 |  |
| 41-50 | 1684 | 18.4 | 5580 | 15.1 | 3727 | 15.6 |  |
| 51-60 | 1883 | 20.5 | 7119 | 19.2 | 5014 | 21.0 |  |
| 61-70 | 1656 | 18.1 | 7842 | 21.2 | 5020 | 21.0 |  |
| 71-80 | 1245 | 13.6 | 6322 | 17.1 | 3808 | 15.9 |  |
| >80 | 808 | 8.8 | 4214 | 11.4 | 2481 | 10.4 |  |
| Total | 9168 | 100.0 | 37001 | 100.0 | 23875 | 100.0 |  |

Source: https://opendatasus.saude.gov.br. The periods were defined based on the epidemiologic week of onset of the first symptoms.

Table 2 compares the occurrence of hospital deaths and ICU use by region and age group between the three defined periods. The North region, which experienced collapse in the provision of healthcare in the first and last periods, emerged in the last, with high percentages of mortality in all age groups, compatible with those observed in the first, in the groups above 60 years, but even greater than those among adults up to 60 years old. It also showed greater use of ICU among patients aged 18-60 and> 80 years. In the Northeast, except for individuals aged 18-30 years, whose mortality remained at a relatively stable level (close to 14%), hospital mortality decreased among adults in the periods, while the percentages of ICU use increased significantly. In the Southeast, there was a decrease in hospital mortality throughout the year for patients aged 60 years and over, and an increase, after falling in the second period, among younger adults, with the highest mortality among those aged 18-30 years. The percentages of ICU use did not vary significantly in most age groups, with an increase in use among patients aged 31-40 years and a reduction between patients aged 71-80 and over 80 years being significant. In the South, the percentages of ICU use dropped significantly, over the periods, among adults over 40 years old, maintaining statistical stability among adults aged 18-40 years. The highest percentage of mortality in the third period among adults aged 18-70 years is highlighted reflecting the severity of the pandemic in the region. In Central West, there were statistically non-significant variations in hospital mortality for adults between 18 and 50 years old, and a drop among adults over 50 years old, with the pattern of ICU use being higher in the third period than in the second, but lower than that in the first period.

**Table 2.** **Covid-19 hospitalization discharges (N=720,363), and comparison of hospital mortality and ICU use by age groups among three periods of the pandemic’s first year in the regions of Brazil**

| Reg./<br>Age<br>group<br>(Ys) | Epidemiologic weeks<br>8/2020 – 25/2020 | | | | | Epidemiologic weeks<br>26/2020 – 43/2020 | | | | | Epidemiologic weeks<br>44/2020 – 7/2021 | | | | | $\chi^2$<br>(p)<br>(1) | $\chi^2$<br>(p)<br>(2) |
| --- | --- | --- | --- | --- | --- | --- | --- | --- | --- | --- | --- | --- | --- | --- | --- | --- | --- |
|  | (16/02/2020 – 20/06/2020) |  |  |  |  | (21/06/2020 – 24/10/2020) |  |  |  |  | (25/10/2020 – 20/02/2021) |  |  |  |  |  |  |
|  | N | Inpatient<br>deaths |  | ICU use |  | N | Inpatient<br>deaths |  | ICU use |  | N | Inpatient<br>deaths |  | ICU use |  |  |  |
|  |  | N | % | N | % |  | N | % | N | % |  | N | % | N | % |  |  |
| North |  |  |  |  |  |  |  |  |  |  |  |  |  |  |  |  |  |
| 0-5 | 335 | 49 | 14,6 | 70 | 20,9 | 736 | 40 | 5,4 | 87 | 11,8 | 645 | 59 | 9,1 | 97 | 15,0 | 0,00 | 0,00 |
| 6-17 | 324 | 41 | 12,7 | 46 | 14,2 | 450 | 28 | 6,2 | 52 | 11,6 | 236 | 24 | 10,2 | 32 | 13,6 | 0,01 | 0,52 |
| 18-30 | 1274 | 159 | 12,5 | 140 | 11,0 | 1200 | 105 | 8,8 | 152 | 12,7 | 1096 | 201 | 18,3 | 202 | 18,4 | 0,00 | 0,00 |
| 31-40 | 2493 | 423 | 17,0 | 408 | 16,4 | 1623 | 175 | 10,8 | 277 | 17,1 | 2451 | 613 | 25,0 | 533 | 21,7 | 0,00 | 0,00 |
| 41-50 | 3323 | 883 | 26,6 | 627 | 18,9 | 2023 | 365 | 18,0 | 404 | 20,0 | 3356 | 1094 | 32,6 | 801 | 23,9 | 0,00 | 0,00 |
| 51-60 | 3815 | 1491 | 39,1 | 958 | 25,1 | 2459 | 653 | 26,6 | 597 | 24,3 | 4073 | 1761 | 43,2 | 1120 | 27,5 | 0,00 | 0,01 |
| 61-70 | 4570 | 2549 | 55,8 | 1346 | 29,5 | 2767 | 1080 | 39,0 | 795 | 28,7 | 4804 | 2687 | 55,9 | 1490 | 31,0 | 0,00 | 0,08 |
| 71-80 | 3879 | 2514 | 64,8 | 1168 | 30,1 | 2420 | 1172 | 48,4 | 786 | 32,5 | 3622 | 2352 | 64,9 | 1171 | 32,3 | 0,00 | 0,06 |
| >80 | 2570 | 1881 | 73,2 | 686 | 26,7 | 1587 | 928 | 58,5 | 465 | 29,3 | 2276 | 1664 | 73,1 | 703 | 30,9 | 0,00 | 0,01 |
| Northeast |  |  |  |  |  |  |  |  |  |  |  |  |  |  |  |  |  |
| 0-5 | 765 | 118 | 15,4 | 183 | 23,9 | 911 | 82 | 9,0 | 254 | 27,9 | 720 | 61 | 8,5 | 225 | 31,3 | 0,00 | 0,01 |
| 6-17 | 507 | 66 | 13,0 | 135 | 26,6 | 655 | 44 | 6,7 | 159 | 24,3 | 334 | 42 | 12,6 | 89 | 26,6 | 0,00 | 0,58 |
| 18-30 | 2647 | 385 | 14,5 | 486 | 18,4 | 1776 | 243 | 13,7 | 413 | 23,3 | 1419 | 192 | 13,5 | 383 | 27,0 | 0,59 | 0,00 |
| 31-40 | 5290 | 919 | 17,4 | 1116 | 21,1 | 3295 | 548 | 16,6 | 852 | 25,9 | 3335 | 492 | 14,8 | 959 | 28,8 | 0,01 | 0,00 |
| 41-50 | 7215 | 1763 | 24,4 | 1694 | 23,5 | 4461 | 972 | 21,8 | 1300 | 29,1 | 4532 | 932 | 20,6 | 1507 | 33,3 | 0,00 | 0,00 |
| 51-60 | 9295 | 3222 | 34,7 | 2681 | 28,8 | 6178 | 1816 | 29,4 | 2033 | 32,9 | 6186 | 1745 | 28,2 | 2242 | 36,2 | 0,00 | 0,00 |
| 61-70 | 10807 | 5159 | 47,7 | 3499 | 32,4 | 7622 | 3028 | 39,7 | 3009 | 39,5 | 7251 | 2721 | 37,5 | 3072 | 42,4 | 0,00 | 0,00 |
| 71-80 | 10273 | 6008 | 58,5 | 3535 | 34,4 | 7897 | 3893 | 49,3 | 3388 | 42,9 | 6636 | 3199 | 48,2 | 3061 | 46,1 | 0,00 | 0,00 |
| >80 | 7814 | 5310 | 68,0 | 2537 | 32,5 | 6733 | 4008 | 59,5 | 2885 | 42,8 | 5312 | 3104 | 58,4 | 2405 | 45,3 | 0,00 | 0,00 |
| Southeast |  |  |  |  |  |  |  |  |  |  |  |  |  |  |  |  |  |
| 0-5 | 909 | 65 | 7,2 | 308 | 33,9 | 1026 | 45 | 4,4 | 283 | 27,6 | 1006 | 44 | 4,4 | 299 | 29,7 | 0,01 | 0,01 |
| 6-17 | 545 | 52 | 9,5 | 158 | 29,0 | 812 | 55 | 6,8 | 257 | 31,7 | 563 | 23 | 4,1 | 167 | 29,7 | 0,00 | 0,53 |
| 18-30 | 4396 | 427 | 9,7 | 1071 | 24,4 | 4368 | 366 | 8,4 | 1103 | 25,3 | 3948 | 424 | 10,7 | 1002 | 25,4 | 0,00 | 0,50 |
| 31-40 | 11624 | 1327 | 11,4 | 3113 | 26,8 | 10643 | 995 | 9,3 | 2853 | 26,8 | 10691 | 1177 | 11,0 | 3006 | 28,1 | 0,00 | 0,04 |
| 41-50 | 16838 | 2815 | 16,7 | 4833 | 28,7 | 16069 | 2236 | 13,9 | 4724 | 29,4 | 16848 | 2421 | 14,4 | 5006 | 29,7 | 0,00 | 0,11 |
| 51-60 | 20977 | 5548 | 26,4 | 6925 | 33,0 | 22077 | 4733 | 21,4 | 7287 | 33,0 | 24043 | 5387 | 22,4 | 7974 | 33,2 | 0,00 | 0,92 |
| 61-70 | 21710 | 8834 | 40,7 | 8053 | 37,1 | 26056 | 8898 | 34,1 | 9831 | 37,7 | 29455 | 10090 | 34,3 | 10973 | 37,3 | 0,00 | 0,31 |
| 71-80 | 16949 | 8994 | 53,1 | 7013 | 41,4 | 21713 | 10050 | 46,3 | 8923 | 41,1 | 24464 | 11052 | 45,2 | 9838 | 40,2 | 0,00 | 0,04 |
| >80 | 13238 | 8639 | 65,3 | 5602 | 42,3 | 16837 | 9669 | 57,4 | 6885 | 40,9 | 18534 | 10655 | 57,5 | 7388 | 39,9 | 0,00 | 0,00 |
| South |  |  |  |  |  |  |  |  |  |  |  |  |  |  |  |  |  |
| 0-5 | 64 | 2 | 3,1 | 17 | 26,6 | 292 | 11 | 3,8 | 66 | 22,6 | 292 | 19 | 6,5 | 64 | 21,9 | 0,24 | 0,72 |
| 6-17 | 41 | 4 | 9,8 | 17 | 41,5 | 223 | 14 | 6,3 | 53 | 23,8 | 197 | 20 | 10,2 | 44 | 22,3 | 0,33 | 0,03 |
| 18-30 | 555 | 33 | 5,9 | 115 | 20,7 | 1683 | 142 | 8,4 | 388 | 23,1 | 1812 | 165 | 9,1 | 387 | 21,4 | 0,06 | 0,36 |
| 31-40 | 1145 | 93 | 8,1 | 284 | 24,8 | 3962 | 299 | 7,5 | 977 | 24,7 | 4706 | 510 | 10,8 | 1142 | 24,3 | 0,00 | 0,88 |
| 41-50 | 1695 | 192 | 11,3 | 521 | 30,7 | 5958 | 768 | 12,9 | 1719 | 28,9 | 7593 | 1117 | 14,7 | 2002 | 26,4 | 0,00 | 0,00 |
| 51-60 | 2014 | 351 | 17,4 | 718 | 35,7 | 8418 | 1624 | 19,3 | 2843 | 33,8 | 10957 | 2382 | 21,7 | 3350 | 30,6 | 0,00 | 0,00 |
| 61-70 | 1904 | 576 | 30,3 | 816 | 42,9 | 9507 | 3225 | 33,9 | 3929 | 41,3 | 13174 | 4620 | 35,1 | 4951 | 37,6 | 0,00 | 0,00 |
| 71-80 | 1395 | 620 | 44,4 | 657 | 47,1 | 7371 | 3424 | 46,5 | 3212 | 43,6 | 10770 | 5069 | 47,1 | 4412 | 41,0 | 0,16 | 0,00 |
| >80 | 883 | 541 | 61,3 | 394 | 44,6 | 5229 | 3041 | 58,2 | 2003 | 38,3 | 7164 | 4193 | 58,5 | 2448 | 34,2 | 0,22 | 0,00 |
| Central West |  |  |  |  |  |  |  |  |  |  |  |  |  |  |  |  |  |
| 0-5 | 64 | 7 | 10,9 | 20 | 31,3 | 344 | 16 | 4,7 | 98 | 28,5 | 126 | 6 | 4,8 | 31 | 24,6 | 0,12 | 0,58 |
| 6-17 | 47 | 4 | 8,5 | 15 | 31,9 | 222 | 12 | 5,4 | 63 | 28,4 | 101 | 5 | 5,0 | 33 | 32,7 | 0,66 | 0,70 |
| 18-30 | 489 | 37 | 7,6 | 124 | 25,4 | 1486 | 143 | 9,6 | 309 | 20,8 | 888 | 90 | 10,1 | 213 | 24,0 | 0,28 | 0,05 |
| 31-40 | 1226 | 135 | 11,0 | 386 | 31,5 | 3573 | 383 | 10,7 | 855 | 23,9 | 2142 | 201 | 9,4 | 546 | 25,5 | 0,20 | 0,00 |
| 41-50 | 1621 | 243 | 15,0 | 542 | 33,4 | 5310 | 833 | 15,7 | 1386 | 26,1 | 3149 | 464 | 14,7 | 882 | 28,0 | 0,47 | 0,00 |
| 51-60 | 1810 | 422 | 23,3 | 707 | 39,1 | 6777 | 1609 | 23,7 | 2135 | 31,5 | 4220 | 907 | 21,5 | 1415 | 33,5 | 0,02 | 0,00 |
| 61-70 | 1605 | 665 | 41,4 | 704 | 43,9 | 7518 | 2666 | 35,5 | 2763 | 36,8 | 4278 | 1450 | 33,9 | 1635 | 38,2 | 0,00 | 0,00 |
| 71-80 | 1215 | 631 | 51,9 | 605 | 49,8 | 6077 | 2814 | 46,3 | 2557 | 42,1 | 3334 | 1560 | 46,8 | 1538 | 46,1 | 0,00 | 0,00 |
| >80 | 799 | 522 | 65,3 | 410 | 51,3 | 4083 | 2451 | 60,0 | 1862 | 45,6 | 2243 | 1301 | 58,0 | 1058 | 47,2 | 0,00 | 0,01 |

Source: https://opendatasus.saude.gov.br. The periods were defined based on the epidemiologic week of onset of the first symptoms. (1) Inpatient deaths; (2) ICU use

Table 3 should be viewed with caution because of the likelihood of underestimating the length of stay in the ICU, especially for patients who fell ill in the third period. Nevertheless, it presents results that suggest that the length of stay in the ICU by Covid-19 has not been prolonged in the last period.

**Table 3.** **Comparison of ICU length of stay among Covid-19 patients (N=190,833) by region and age group, among three periods of the pandemic’s first year In Brazil**

| Reg./<br>Age<br>group<br>(Ys) | Epidemiologic weeks<br>8/2020-25/2020 |  |  |  |  | Epidemiologic weeks<br>26/2020-43/2020 |  |  |  |  | Epidemiologic weeks<br>44/2020-7/2021 |  |  |  |  | KW<br>Test<br>(p) |
| --- | --- | --- | --- | --- | --- | --- | --- | --- | --- | --- | --- | --- | --- | --- | --- | --- |
|  | (16/02/2020 – 20/06/2020) |  |  |  |  | (21/06/2020 – 24/10/2020) |  |  |  |  | (25/10/2020 – 20/02/2021) |  |  |  |  |  |
|  | N | Mean | Std | Md | Max | N | Mean | Std | Md | Max | N | Mean | Std | Md | Max |  |
| North |  |  |  |  |  |  |  |  |  |  |  |  |  |  |  |  |
| 0-5 | 54 | 13.3 | 26.4 | 6 | 157 | 42 | 10.1 | 14.9 | 5 | 70 | 52 | 11.1 | 15.8 | 5.5 | 92 | 0.90 |
| 6-17 | 36 | 11.2 | 15.2 | 7 | 84 | 32 | 5.7 | 5.5 | 5 | 22 | 19 | 8.9 | 7.3 | 6 | 25 | 0.11 |
| 18-30 | 113 | 11.2 | 10.8 | 8 | 57 | 113 | 11.6 | 12.9 | 7 | 86 | 147 | 9.8 | 8.6 | 7 | 58 | 0.93 |
| 31-40 | 324 | 9.7 | 9.2 | 7 | 50 | 194 | 10.2 | 9.5 | 7 | 63 | 426 | 9.6 | 8.6 | 7 | 46 | 0.88 |
| 41-50 | 529 | 10.5 | 12.0 | 7 | 108 | 318 | 10.9 | 10.2 | 8 | 57 | 636 | 10.8 | 10.3 | 7.5 | 68 | 0.23 |
| 51-60 | 847 | 10.3 | 12.7 | 7 | 130 | 495 | 11.7 | 10.6 | 9 | 71 | 939 | 10.7 | 10.2 | 8 | 107 | 0.00 |
| 61-70 | 1220 | 10.2 | 13.5 | 7 | 223 | 672 | 12.8 | 14.6 | 8 | 146 | 1283 | 10.8 | 9.4 | 8 | 67 | 0.00 |
| 71-80 | 1082 | 9.2 | 10.2 | 6 | 98 | 677 | 11.9 | 18.0 | 7 | 357 | 1042 | 9.5 | 8.4 | 7 | 63 | 0.00 |
| >80 | 625 | 7.7 | 8.7 | 5 | 67 | 418 | 10.2 | 12.1 | 7 | 77 | 641 | 8.5 | 8.1 | 6 | 60 | 0.00 |
| Northeast |  |  |  |  |  |  |  |  |  |  |  |  |  |  |  |  |
| 0-5 | 109 | 11.4 | 15.8 | 5 | 96 | 110 | 12.3 | 16.9 | 6 | 109 | 85 | 11.8 | 15.0 | 5 | 89 | 0.67 |
| 6-17 | 88 | 10.6 | 13.5 | 6 | 70 | 82 | 10.1 | 13.3 | 5.5 | 81 | 47 | 10.9 | 14.0 | 5 | 59 | 0.98 |
| 18-30 | 342 | 10.3 | 16.5 | 5 | 175 | 264 | 10.4 | 16.1 | 5 | 187 | 256 | 8.8 | 10.2 | 6 | 78 | 0.91 |
| 31-40 | 768 | 10.2 | 11.9 | 7 | 105 | 536 | 9.9 | 11.0 | 6.5 | 89 | 588 | 8.6 | 9.4 | 6 | 83 | 0.15 |
| 41-50 | 1265 | 11.0 | 14.1 | 7 | 147 | 876 | 10.3 | 11.8 | 7 | 118 | 998 | 9.1 | 9.0 | 7 | 98 | 0.81 |
| 51-60 | 2094 | 12.0 | 14.0 | 8 | 149 | 1388 | 11.4 | 12.4 | 8 | 98 | 1528 | 11.0 | 9.9 | 8 | 68 | 0.42 |
| 61-70 | 2879 | 11.3 | 13.1 | 8 | 176 | 2247 | 11.9 | 12.9 | 8 | 146 | 2221 | 11.0 | 10.2 | 8 | 102 | 0.07 |
| 71-80 | 3062 | 11.0 | 12.7 | 7 | 155 | 2679 | 11.9 | 13.2 | 8 | 161 | 2319 | 11.0 | 10.2 | 9 | 93 | 0.00 |
| >80 | 2217 | 9.8 | 13.3 | 6 | 276 | 2352 | 9.6 | 10.7 | 7 | 155 | 1943 | 9.4 | 8.8 | 7 | 75 | 0.02 |
| Southeast |  |  |  |  |  |  |  |  |  |  |  |  |  |  |  |  |
| 0-5 | 157 | 13.4 | 24.1 | 6 | 187 | 141 | 10.3 | 15.9 | 5 | 103 | 123 | 7.9 | 9.8 | 4 | 49 | 0.10 |
| 6-17 | 100 | 7.7 | 9.7 | 4 | 62 | 144 | 10.4 | 20.3 | 5 | 190 | 71 | 8.4 | 9.3 | 6 | 55 | 0.78 |
| 18-30 | 690 | 9.5 | 12.7 | 6 | 183 | 665 | 9.4 | 11.7 | 5 | 108 | 611 | 8.6 | 9.5 | 5 | 55 | 0.10 |
| 31-40 | 1923 | 10.1 | 12.3 | 6 | 151 | 1765 | 10.1 | 12.5 | 6 | 164 | 1792 | 9.2 | 9.3 | 6 | 83 | 0.78 |
| 41-50 | 3110 | 10.8 | 12.3 | 7 | 204 | 3005 | 11.3 | 13.0 | 7 | 182 | 3128 | 9.8 | 11.3 | 7 | 374 | 0.64 |
| 51-60 | 4942 | 12.6 | 14.3 | 9 | 225 | 5062 | 13.1 | 14.5 | 9 | 212 | 5393 | 11.4 | 10.6 | 8 | 108 | 0.76 |
| 61-70 | 6330 | 13.4 | 15.9 | 9 | 289 | 7511 | 13.4 | 13.7 | 9 | 164 | 8059 | 12.1 | 10.9 | 9 | 97 | 0.06 |
| 71-80 | 5807 | 12.6 | 15.1 | 8 | 240 | 7291 | 12.7 | 13.1 | 9 | 174 | 7615 | 11.8 | 10.4 | 9 | 97 | 0.00 |
| >80 | 4783 | 11.3 | 14.4 | 7 | 209 | 5806 | 10.3 | 11.5 | 7 | 191 | 6003 | 10.0 | 9.2 | 8 | 102 | 0.02 |
| South |  |  |  |  |  |  |  |  |  |  |  |  |  |  |  |  |
| 0-5 | 16 | 11.7 | 14.3 | 5.5 | 44 | 58 | 15.5 | 23.1 | 7.5 | 129 | 49 | 10.3 | 9.5 | 7 | 43 | 0.73 |
| 6-17 | 15 | 18.5 | 13.7 | 19 | 53 | 47 | 12.9 | 14.5 | 6 | 61 | 33 | 10.5 | 12.4 | 7 | 58 | 0.09 |
| 18-30 | 103 | 11.0 | 12.8 | 6 | 83 | 352 | 10.0 | 10.6 | 7 | 71 | 321 | 9.8 | 10.8 | 7 | 65 | 0.97 |
| 31-40 | 255 | 12.3 | 12.2 | 8 | 85 | 859 | 12.0 | 14.0 | 7 | 110 | 946 | 10.4 | 10.0 | 7 | 68 | 0.27 |
| 41-50 | 477 | 12.2 | 13.2 | 8 | 108 | 1575 | 12.9 | 13.8 | 9 | 177 | 1727 | 11.7 | 11.2 | 8 | 105 | 0.17 |
| 51-60 | 661 | 14.4 | 14.7 | 9 | 105 | 2600 | 14.3 | 14.9 | 10 | 169 | 2955 | 12.7 | 11.0 | 10 | 99 | 0.55 |
| 61-70 | 761 | 15.5 | 14.7 | 11 | 120 | 3693 | 14.9 | 14.9 | 11 | 138 | 4577 | 13.2 | 11.5 | 10 | 97 | 0.00 |
| 71-80 | 631 | 14.8 | 16.9 | 10 | 222 | 3086 | 13.9 | 13.9 | 10 | 156 | 4170 | 12.3 | 11.1 | 9 | 105 | 0.00 |
| >80 | 384 | 11.6 | 12.0 | 8 | 89 | 1933 | 10.2 | 11.7 | 7 | 185 | 2335 | 9.7 | 9.1 | 7 | 95 | 0.01 |
| Central West |  |  |  |  |  |  |  |  |  |  |  |  |  |  |  |  |
| 0-5 | 13 | 7.4 | 7.4 | 3 | 20 | 63 | 7.9 | 10.6 | 4 | 63 | 27 | 8.7 | 10.0 | 5 | 40 | 0.64 |
| 6-17 | 6 | 6.8 | 7.3 | 4.5 | 20 | 40 | 9.7 | 9.9 | 6 | 46 | 26 | 8.0 | 8.7 | 4 | 33 | 0.44 |
| 18-30 | 85 | 8.8 | 10.4 | 4 | 47 | 198 | 10.2 | 12.9 | 6 | 110 | 156 | 7.7 | 6.8 | 5 | 42 | 0.38 |
| 31-40 | 252 | 11.4 | 16.7 | 6.5 | 177 | 538 | 9.3 | 9.7 | 6 | 53 | 368 | 9.1 | 8.5 | 7 | 72 | 0.64 |
| 41-50 | 357 | 11.2 | 12.5 | 7 | 112 | 972 | 11.2 | 12.9 | 7.5 | 125 | 649 | 10.4 | 10.5 | 7 | 96 | 0.85 |
| 51-60 | 499 | 12.5 | 16.1 | 8 | 202 | 1572 | 12.5 | 14.3 | 8 | 163 | 1063 | 11.3 | 9.9 | 9 | 83 | 0.46 |
| 61-70 | 580 | 13.5 | 14.1 | 9 | 112 | 2192 | 12.7 | 13.7 | 9 | 159 | 1308 | 11.9 | 10.0 | 9 | 89 | 0.62 |
| 71-80 | 510 | 13.4 | 14.0 | 9 | 99 | 2137 | 11.8 | 12.7 | 8 | 156 | 1317 | 12.0 | 10.6 | 9 | 85 | 0.03 |
| >80 | 366 | 11.2 | 12.4 | 8 | 112 | 1637 | 10.3 | 11.8 | 7 | 212 | 937 | 9.8 | 8.8 | 7 | 66 | 0.66 |

Source: https://opendatasus.saude.gov.br. The periods were defined based on the epidemiologic week of onset of the first symptoms. Std=Standard deviation; Md=Median; Max=Maximum. KW=Kruskal-Wallis Test.

## Final considerations

Considering the analyses undertaken, based on data referring to February 20, 2021, this study does not provide evidence to support a greater participation of young adults in hospitalizations in the third period. However, the differentiated rise in Covid-19 hospital deaths among adults aged 18-50 and even 50-60 years in the North, compared especially to the first period, also marked by a collapse in the health system in some states, is a warning which meets the hypothesis of greater severity of the disease caused by the P1 variant in this population. In any case, the results showed that the participation of young adults in hospitalizations for Covid-19 was never negligible and was always related to hospital mortality rates close to or above 10%. In this study, the extension of the ICU use time was not observed.

The risk of an increase in severe cases among adults up to 50 years of age is open to multifactorial explanations and confronts a certain common sense that the disease would offer a low risk for younger people. The Covid-19 “youthening” phenomenon in Brazil is based on the country’s own socio-demographic and economic characteristics and may have been strengthened by the increasing circulation of viral variants. It is important to continue monitoring its progression and effects. Analyses from SIVEP itself and from the Civil Registry are beginning to point out the rapid growth of Covid-19 cases and deaths in individuals between 20 and 59 years old since the beginning of the year^8,9^. The focus on hospitalizations is more affected by data delays. The results, however, provide a baseline. Explorations at less aggregate levels, in capitals and large cities, which suffer markedly from the effects of large agglomerations, may be able to add more specific and circumscribed findings about the rejuvenation of the pandemic reported by health and media professionals^8,9^.

## Data Availability

The manuscript is based on data publicly available.

https://opendatasus.saude.gov.br

